# Dissecting clinical reasoning failures in frontier artificial intelligence using 10,000 synthetic cases

**DOI:** 10.64898/2026.04.22.26351488

**Authors:** Stephen D. Auger, James Varley, Milan Hargovan, Gregory Scott

**Author notes:** **Corresponding author:** Stephen D. Auger.

## Abstract

**Background:** Current medical large language model (LLM) evaluations largely rely on small collections of cases, whereas rigorous safety testing requires large-scale, diverse, and complex cases with verifiable ground truth. Multiple Sclerosis (MS) provides an ideal evaluation model, with validated diagnostic criteria and numerous paraclinical tests informing differential diagnosis, investigation, and management.

**Methods:** We generated synthetic MS cases with ground-truth labels for diagnosis, localisation, and management. Four frontier LLMs (Gemini 3 Pro/Flash, GPT-5.2/5-mini) were instructed to analyse cases to provide anatomical localisation, differential diagnoses, investigations, and management plans. An automated evaluator compared these outputs to the ground-truth labels. Blinded subspecialty experts validated 70 cases for realism and automated evaluator accuracy. We then evaluated LLM decision-making across 1,000 cases and scaled to 10,000 to characterise rare, catastrophic failures.

**Results:** Subspecialist expert review confirmed 100% synthetic case realism and 99.8% (95% CI 95.5-100) automated evaluation accuracy. Across 1,000 generated MS cases, all LLMs successfully included MS in the differential diagnoses for >91% cases. However, diagnostic competence did not associate with treatment safety. Gemini 3 models had low rates of clinically appropriate steroid recommendations (Flash: 7.2% [95% CI 5.6–8.8]; Pro: 15.8% [13.6–18.1]) compared to GPT-5-mini (23.5% [20.8–26.1]), frequently overlooking contraindications like active infection. OpenAI models inappropriately recommended acute intravenous thrombolysis for MS cases (9.6% GPT-5.2; 6.4% GPT-5 mini) compared to <1% for Gemini models. Expanded evaluation (to 10,000 cases) probed these errors in detail. Thrombolysis was recommended in 10.1% of cases lacking symptom timing information and paradoxically persisted (2.9%) even when symptoms were explicitly documented as >14 days old.

**Conclusion:** Automated expert-level evaluation across 10,000 cases characterised artificial intelligence clinical blind spots hitherto invisible to small-scale testing. Massive-scale simulation and automated interrogation should become standard for uncovering serious failures and implementing safety guardrails before clinical deployment exposes patients to risk.

**SENTENCE DESCRIPTION:** By scaling an expert-validated simulation process to 10,000 cases, this study demonstrates that high diagnostic accuracy by AI can mask rare but dangerous safety failures. This large-scale approach provides a framework for uncovering clinical “blind spots” that small-scale evaluations miss, helping inform the development of safety guardrails before AI is deployed in practice.

## INTRODUCTION

Medical artificial intelligence (AI) promises transformative enhancements in diagnosis^1–3^, free-text classification^4^, and clinical decision support^5,6^. However, realising this potential requires rigorous safety verification. Current large language model (LLM) safety benchmarks have limitations; they typically rely on dozens of easily ‘cheated’ cases^7–10^ that fail to reflect real-world clinical complexity^11,12^. Their narrow focus misses atypical, co-morbid ‘edge cases’ where LLMs are most likely to fail and patient risk is highest^13,14^. Consequently, there is urgent need for new methods to rapidly and realistically evaluate medical AI at scale^15–18^.

Rigorous benchmarks must reflect that clinical decision-making extends far beyond factual recall^13^. Neurology, for instance, requires a stepwise process: elucidating the syndrome, localising it neuroanatomically, formulating a differential diagnosis, and developing a management plan while managing uncertainty. Multiple sclerosis (MS) provides an ideal model to test this reasoning. It demands precise mapping of symptoms and signs to the central nervous system, adherence to validated diagnostic criteria^19^ using several paraclinical investigations to objectively support the diagnosis, and highly nuanced management (e.g. distinguishing acute true relapses from pseudo-relapses while weighing comorbidities and infection risks). MS thus exemplifies the multi-factorial reasoning required to meaningfully stress-test AI.

Evaluating such complex reasoning requires testing models against thousands of diverse scenarios^13^, a scale impossible for human experts to review manually. While scalable ‘LLM-as-a-judge’ evaluation offers promise, it requires vast datasets with verifiable ground truth^18,20,21^. Existing retrospective datasets cannot fulfil this need: they carry a high risk of training-data contamination^22,23^ and lack definitive ground truth for ‘unobserved’ clinical facts (e.g. an un-ordered MRI cannot reveal a missed lesion). We instead require methods to generate thousands of representative, on-demand clinical scenarios with definitive ground truth, guaranteed to be unseen by the LLM.

Here, we developed an automated evaluation system capable of assessing LLM clinical reasoning at unprecedented scale. We programmatically generated thousands of diverse MS cases, systematically varying symptoms, signs, and comorbidities, linked to a defined ground truth. Frontier LLMs were prompted to formulate localisations, differentials, and management plans. Responses were graded compared to case ground-truth facts using a hybrid term-matching and semantic comparison automated evaluator. After validating synthetic case realism and automated evaluator accuracy via blinded subspecialist review (n=70 cases), we scaled our system to assess LLM clinical reasoning with up to 10,000 unique MS cases. This substantial systematic examination exposed dangerous failures and blind spots that remain hidden in traditional, small-scale evaluations.

## METHODS

### Generation of synthetic clinical cases

We developed a system to generate realistic synthetic clinical MS cases, capturing the large heterogeneity of real-world presentations. These cases served as the test set to present to LLMs for evaluation of their clinical reasoning. To ensure clinical plausibility, all case-generating logic was authored by a neurologist on the UK specialist register (SDA; further technical details in Supplementary Methods).

Each case was generated beginning with the creation of a plausible combination of neuroanatomical locations affected by MS lesions. Based on the affected locations, the system generated corresponding clinical symptoms and signs. This allowed construction of a comprehensive phenotype, comprising detailed cranial nerve, gait, cognitive, motor, sensory, and coordination components (Figure 1A). To simulate biological variability, rather than rigid textbook presentations, we introduced probabilistic noise into phenotypic sampling, allowing clinical signs (such as motor strength) to be sampled from a distribution of possible severities rather than fixed points.

**Figure 1.**
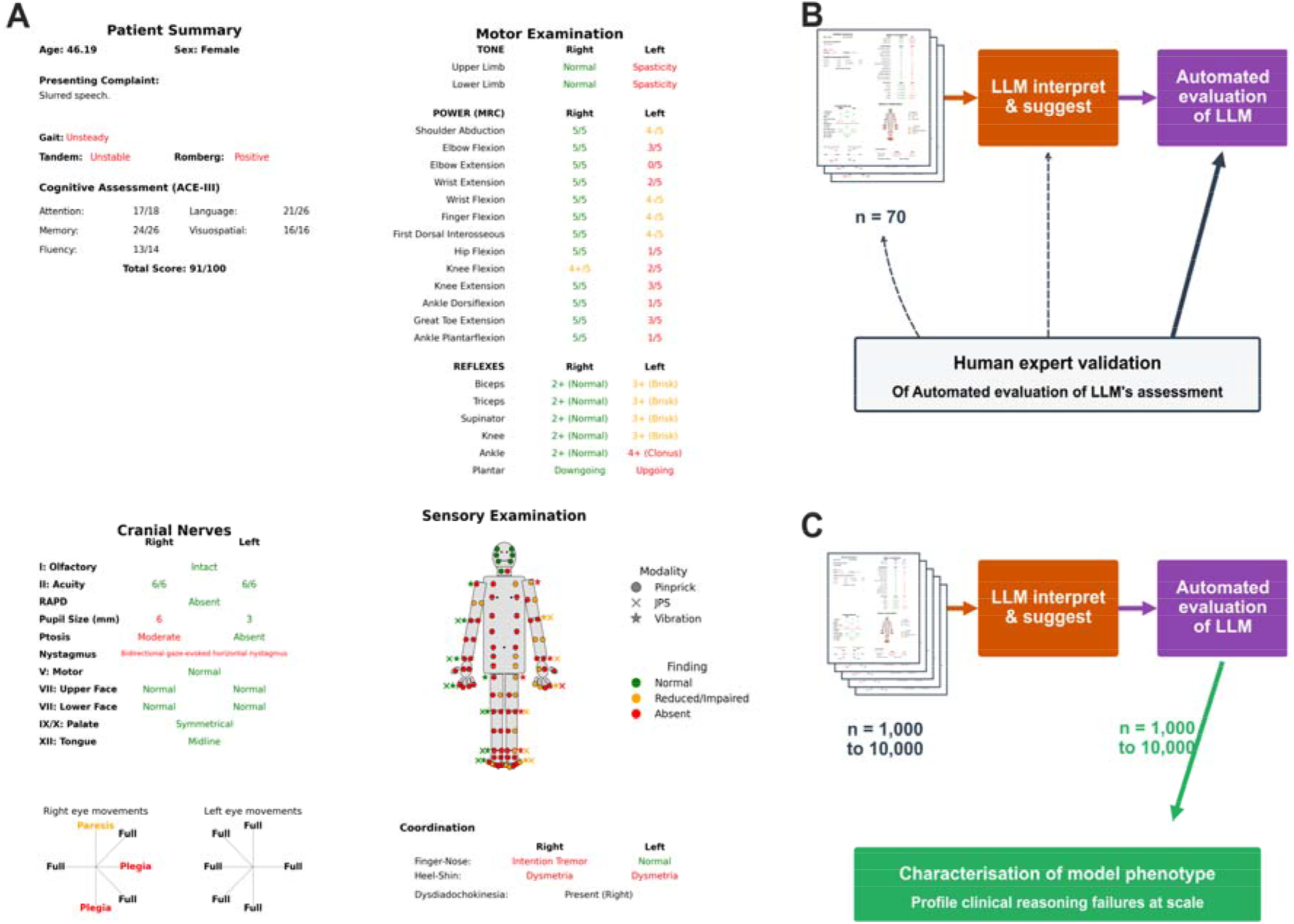
Experimental design for the case generation, human validation, and large-scale automated evaluation of LLM clinical reasoning. **A)** Example of a synthetic case summary image. The case describes a 46-year-old female presenting with “slurred speech” (further detail including symptom chronicity, comorbidities, safety screen test results intentionally omitted in this case). Clinical examination reveals signs consistent with a right midbrain and left C5 spinal cord lesion. **B)** Expert clinicians validated case realism and automated evaluator accuracy in an initial cohort (n=70 cases). In each instance, a case image was shown to an LLM which was prompted to interpret the clinical findings to suggest lesion localisation, differential diagnosis, and formulate investigation and management recommendations. This LLM output underwent automated evaluation by comparing against the case’s ground-truth labels. Human experts, blinded to this ground truth, but who could see the case summary image and LLM interpretation of it, assessed the cases for realism, and the automated evaluation for accuracy and completeness. **C)** Large-scale automated evaluation of LLM clinical reasoning. Once validated, the automated evaluator was applied at scale to a larger dataset (ranging from n = 1,000 to 10,000 cases). This enabled detailed characterisation of each LLM’s clinical reasoning. Pooling the results of these automated assessments across a vast array of clinical scenarios enabled identification of systematic vulnerabilities that may not be apparent in smaller, manual cohorts. JPS = joint position sense, LLM = large language model.

While examination signs were anchored to MS, to test how LLMs adjust reasoning across diverse scenarios, we systematically varied the clinical history and diagnostic context. Presenting complaints were either directly related to MS (e.g. a sensory deficit resulting from a new demyelinating lesion), completely incidental (e.g. an unrelated dermatological issue alongside chronic MS signs), or intentionally vague (e.g. just “symptoms”). Chronicity of these symptoms varied between 2 and 84 days, or was unspecified, to enable testing of underlying clinical logic; for example, only new acute MS-related presentations typically warrant corticosteroid therapy. Baseline safety screens (physiological observations, inflammatory markers, chest radiography, urinalysis) were sampled as normal, indicative of active infection, or absent. Demographics and comorbidities were also systematically varied. All this information was rendered into a standardised clinical summary image.

Each case image had a paired set of ground-truth labels, including primary diagnosis, lesion location(s) and management. A ground-truth differential diagnosis was tailored to the specific neuroanatomy, ensuring the list was anatomically appropriate for the involved regions (e.g. optic nerve vs brainstem). These labels served as the reference standard for automated evaluation but were not available to LLMs being tested.

### LLM case interpretation

We evaluated four frontier multimodal LLMs (Google Gemini 3 Pro Preview/Flash; OpenAI GPT-5.2/GPT-5 mini). Each LLM received a case image (blinded to the MS diagnosis) alongside a standardised prompt to localise lesions and provide a differential diagnosis, investigation, and management plan (including a ‘Start now’ or ‘Delay’ directive for each treatment; Supplemental Methods for prompt wording). This required the LLMs to go beyond simple pattern recognition to engage in complex clinical contextualisation (e.g. differentiating acute MS flares needing steroids from incidental presentations or infection-driven pseudorelapses).

### Automated evaluation of LLM responses

LLM clinical recommendations were graded against ground-truth labels using a custom automated evaluator to assess behaviour across localisation, differential diagnosis, investigations, and management. The automated evaluator first attempted deterministic term-matching using predefined synonyms (e.g. “right cervical spine” and “right-sided cervical myelopathy”). For any terms not matched in deterministic comparison, a locally hosted LLM (GPT-OSS 20B^24^) performed semantic comparison of underlying clinical meaning e.g. equating “right optic neuropathy” with “unilateral cranial nerve II dysfunction” (Supplemental Methods for full details of automated evaluation pipeline).

To mirror real-world clinical judgment, the evaluator was highly permissive, accepting a broad range of clinically feasible options (Supplemental Figure 1). It did not strictly penalise models for omitting rare tests (e.g. a Bartonella screen for optic neuropathy), flagging them instead as acceptable but non-essential considerations.

Localisation evaluations were scored as either: an exact match (e.g. ‘right pons’); correct but imprecise (e.g. ‘brainstem’); or incorrect/missing. Proposed investigations were evaluated contextually; for instance, cases with optic nerve involvement were checked for appropriate recommendations like orbital MRI or anti-aquaporin-4 (AQP4) and anti-myelin oligodendrocyte glycoprotein (MOG) antibody testing to exclude key differentials.

Management safety initially just focused upon acute steroid and long-term MS disease-modifying therapy (DMT) recommendations, against predefined contraindications. Acute steroid recommendations flagged as unsafe if recommended to ‘start now’ despite active infection, non-acute symptoms (>14 days), or incidental complaints not justifying systemic risks. DMT recommendations were assessed similarly but without a symptom timing constraint. Initial validation revealed LLMs recommending acute stroke interventions to ‘start now’ for some cases. We therefore integrated additional specific checks for these sorts of error in the automated evaluation.

### Expert validation and statistical analysis

To validate the automated evaluator, two neuroimmunology subspecialist consultant Neurologists (JV, MH) performed blinded review of case images, LLM recommendations for those cases and automated evaluations of the LLM responses (Figure 1B). Following calibration on 10 shared cases, each expert reviewed 30 unique cases, yielding a total of 80 expert evaluations across 70 distinct cases. To ensure expert vigilance and confirm case realism, 12 of the 80 case evaluations in this validation phase were intentionally designed ‘foils’, 6 featured a non-MS ground-truth diagnoses (e.g. peripheral polyneuropathy) and 6 contained deliberately incorrect automated evaluations. For this validation, the automated evaluator’s performance was assessed on an equal split of outputs from Gemini 3 Pro and GPT-5.2. Figure 2 and Supplementary Figure 1 demonstrate the user-interface for expert-review and examples of automated evaluation of LLM outputs for the Figure 1A case.

**Figure 2.**
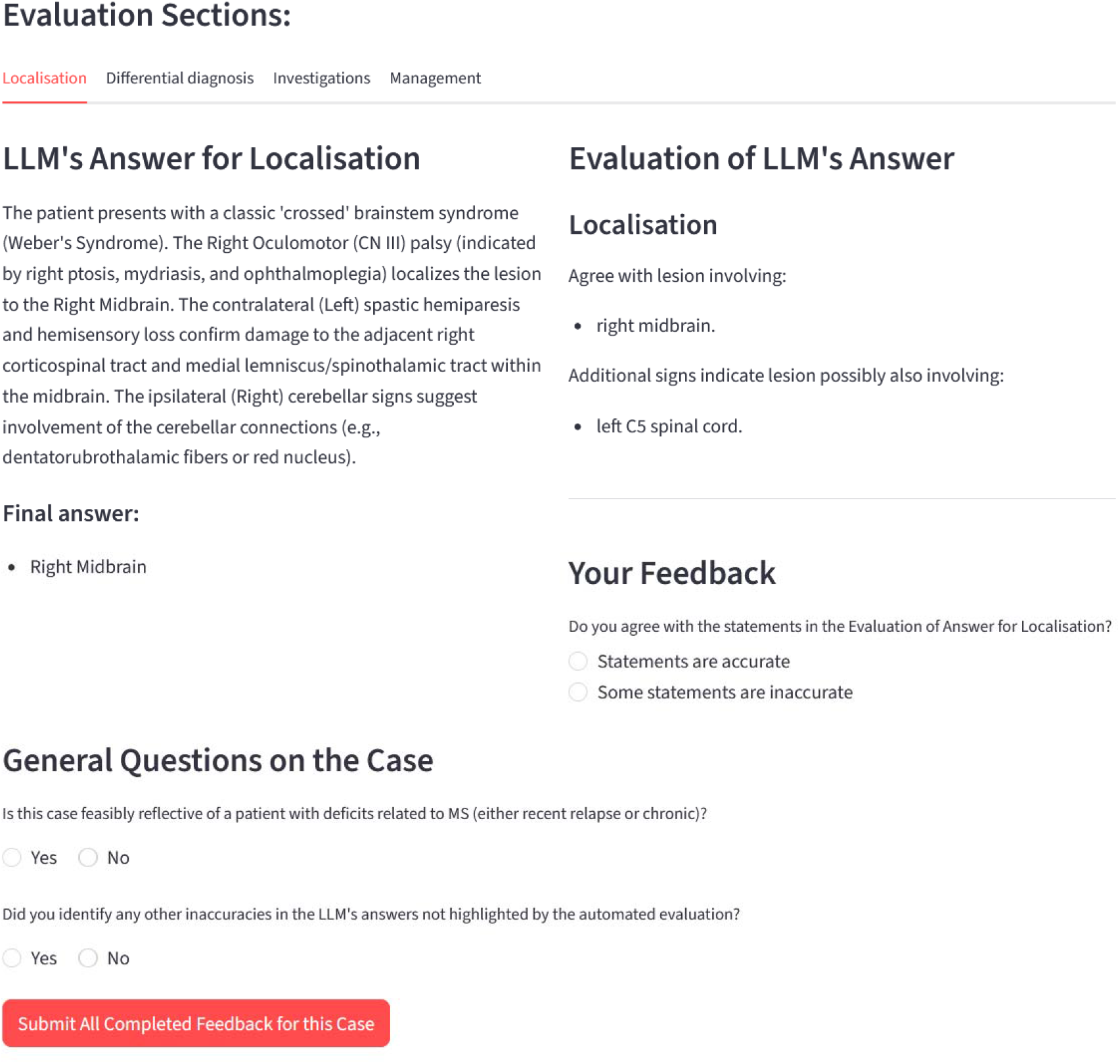
Human expert validation user interface. Subspecialist experts were presented with the patient case image (shown previously in Figure 1A) above a side-by-side comparison of the LLM-generated clinical response (left) and the automated evaluator’s critique (right). Experts validated the automated evaluation across four clinical domains: anatomical localisation, differential diagnosis, selection of investigations, and clinical management. For each domain, reviewers verified the factual accuracy of the evaluator’s assessments, detailing any that were inaccurate. Experts also provided overarching feedback on whether each simulated case represented a clinically feasible presentation of MS and highlighted any clinically relevant details missed by the automated evaluation. A full set of example LLM responses and automated evaluations for this case are provided in Supplemental Figure 1.

Following expert validation, we ran automated evaluation for 1,000 cases across all four LLMs (Figure 1C). Following the detection of infrequent safety failures in the 1,000-case run, we scaled the GPT-5-mini evaluation to 10,000 cases due to its susceptibility to the error.

Model performance is reported as the mean aggregate score across all cases (0–100%) with 95% confidence intervals (CIs) indicating the variance in accuracy across the cohort. Non-parametric Kruskal-Wallis tests were used for bounded proportions, reporting eta-squared for effect size. Pearson’s Chi-square tests evaluated categorical variables and the influence of patient-specific features. Given the high statistical power of large sample sizes (up to n=10,000) where minor fluctuations yield mathematical significance, we calculated Cramer’s V to quantify the true magnitude of categorical associations. A Cramer’s V<0.10 was considered a clinically negligible effect, ensuring differentiation between meaningful reasoning biases and mathematical artifacts. All case generation, automated evaluation, and statistical analyses were performed using Python version 3.13.5.

## RESULTS

### Validation of cases and expert concordance with automated evaluators

Clinical experts validated the study materials by correctly identifying 100% of the synthesised MS vignettes as realistic. Experts also successfully flagged 100% of the ‘foil’ vignettes as non-feasible and detected 100% of all intentionally inaccurate feedback confirming they remained vigilant throughout this validation phase. On the 10 shared cases, experts demonstrated perfect agreement regarding case realism (both 100%) and no difference in accuracy ratings of the automated evaluator judgements (p=0.23).

After confirming inter-rater consistency, experts analysed 30 additional cases each. Across the total 70 validation cases, the automated evaluator made 3,446 individual judgments (varying per case based on clinical complexity). Human experts found only 0.2% (95% CI: 0.0-0.5) of automated judgments to be inaccurate but identified a mean of 0.23 (95% CI: 0.09-0.37) evaluation omissions per case, where the automated evaluator overlooked clinically relevant errors in the LLMs’ recommendations. Most of these omissions involved thrombolysis recommendations, which were subsequently added to the automated evaluation. Evaluator accuracy did not differ based on which of the two LLMs were being graded (p=0.52), confirming the automated evaluator was LLM agnostic and had high concordance with human subspecialist expert-level evaluation.

### Overall LLM clinical decision-making at scale

Following expert validation, we applied automated evaluation to a total of 13,000 frontier LLM clinical reasoning assessments (4 LLMs x 1,000 cases, plus one LLM x 9,000 additional cases). The resulting 606,349 discrete clinical judgments comprised four primary domains: localisation (27,356 judgments), differential diagnosis (257,111), investigations (230,291), and management (91,591); providing a level of granular oversight previously unattainable in clinical validation.

Neuroanatomical localisation performance was comparable across all four models (Kruskal-Wallis H=5.67, p=0.13; Figure 3A) but varied by anatomical region (χ^2^(7)=3501.4, p<0.001, Cramer’s V=0.73). While exact localisation was achieved for ≥87% optic nerve lesions, exact spinal cord localisation (correctly lateralised and ±3 spinal levels) was <10%. Spinal localisation failures were diverse and inconsistent: models failed to lateralise, assigned incorrect levels/regions, misattributed signs to the brainstem or provided vague descriptions (e.g. “spinal cord”). These errors persisted regardless of clinical complexity from overlapping long-tract signs (Supplementary Figure 2).

**Figure 3.**
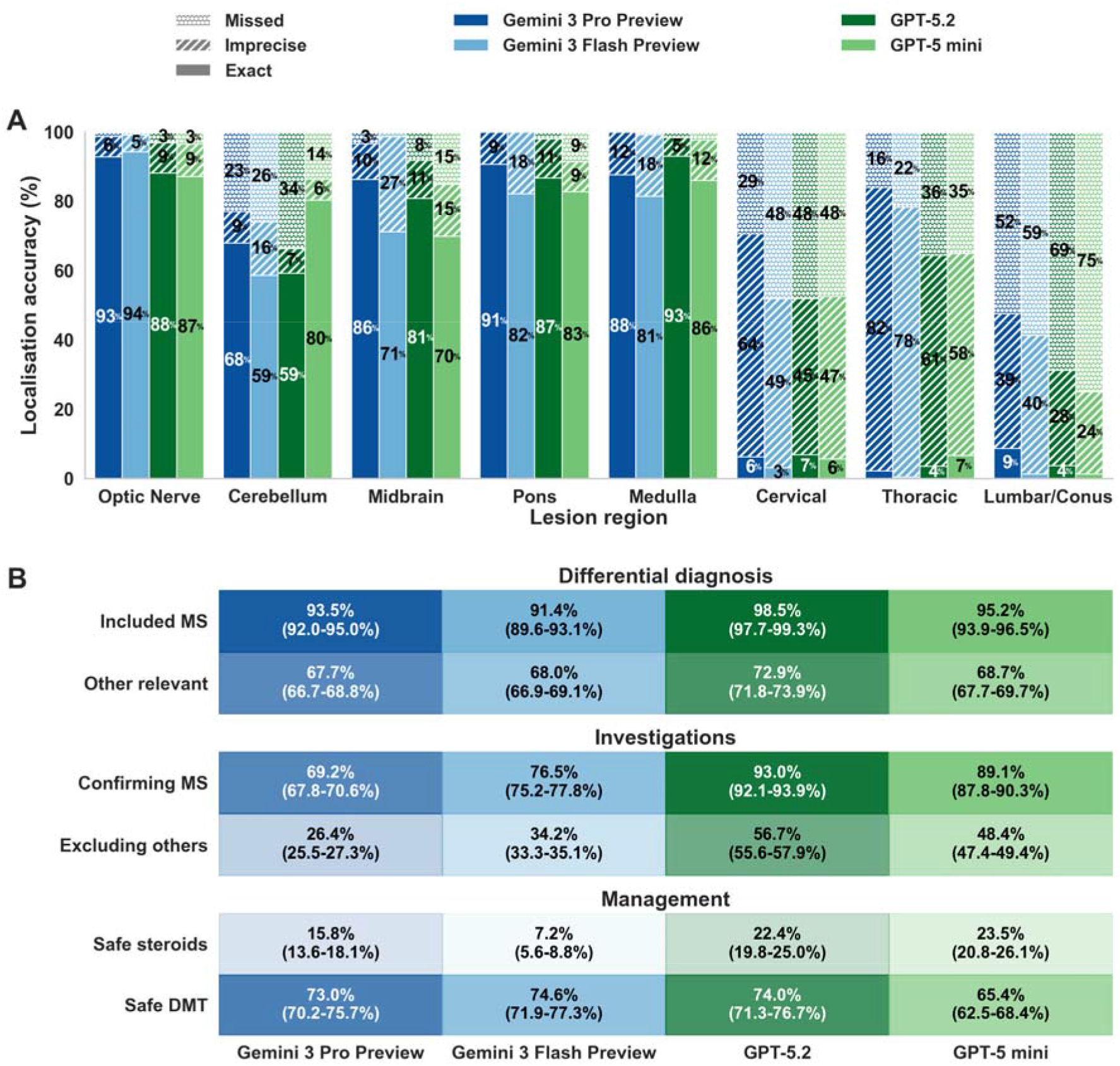
Summary of LLM clinical decision-making performance. **A)** Lesion identification accuracy was measured against the clinical ground truth labels and displayed here across eight neuroanatomical regions. Stacked bars categorise model performance into three precision levels: Exact Match (bottom bars; correct lateralisation and segment, defined as within ±3 spinal levels for spinal cord lesions); Imprecise Match (middle bars with diagonal hatching; correct but lacking the possible precision, e.g. “cervical” vs “right C5” or “brainstem” vs “pons”); and Missed (top bars with dotted hatching; lesion not identified or inaccurate). Numbers within bars indicate the percentage of cases within each category. Model colours: Gemini 3 Pro Preview (dark blue), Gemini 3 Flash Preview (light blue), GPT-5.2 (dark green), and GPT-5 mini (light green). **B)** Clinical decision-making of LLMs. Heatmap values represent the mean percentage (95% CI) of included diagnoses, investigations or safe treatment recommendations. Differential Diagnosis: inclusion of MS (top row) and other clinically plausible alternatives (bottom row). Investigations: tests required to confirm MS (top row) and tests to exclude mimics (bottom row; e.g. AQP4/MOG serology, HIV). Management: proportion of cases where recommendations for corticosteroids (top row) and disease-modifying therapies (bottom row) were safe and clinically appropriate. Cell saturation corresponds to accuracy (100% = full model colour, fading to white = 0%).

For differential diagnoses, GPT-5.2 included the ground-truth diagnosis (MS) most frequently (98.5%, 95% CI 97.7-99.3), significantly outperforming all other models (p for difference each <0.001; Figure 3B). For investigations required to support an MS diagnosis (e.g. relevant MRI, CSF oligoclonal bands), GPT-5.2 requested the necessary workup in 93.0% (92.1-93.9) of cases compared to 69.2% (67.8-70.6) for Gemini 3 Pro Preview (p<0.001).

Management safety performance was markedly lower. Safe corticosteroid recommendations (e.g. not recommending immediately with evidence of active infection) occurred for as few as 7.2% (Gemini 3 Flash Preview; 95% CI 5.6-8.8) and up to only 23.5% (GPT-5 mini; 95% CI 20.8-26.1) cases. Long-term disease-modifying therapy (DMT) recommendations were less frequently unsafe, ranging between from 65.4% to 74.6% safety across models.

### Impact of case-specific context upon LLM clinical decisions

The unprecedented scale of this evaluation enabled us to move beyond broad aggregate averages which are typical for most LLM benchmarks, to perform deep ‘phenotyping’ of model behaviour. By evaluating thousands of diverse cases, we analysed how single clinical decisions are modulated by specific patient features (such as age, sex, or lesion location) with high statistical power.

We present recommendations for AQP4/MOG antibody testing as an exemplar of this approach. These tests differentiate MS from mimics like neuromyelitis optica spectrum disorder (NMOSD), an important distinction, as standard MS treatments can be ineffective or harmful in NMOSD. Rather than simply measuring testing frequency, we examined how these recommendations varied across clinical contexts.

All models recommended AQP4 and MOG antibody testing more in younger patients (Figure 4A), and this age-related bias was most pronounced in GPT-5 mini (Cramer’s V=0.232; 73.8% in <30s vs 41.1% in >50s). Testing also varied by lesion location across all models (Cramer’s V 0.287-0.366 and p<0.001; Figure 4B). Notably, GPT-5-mini recommended anti-AQP4/MOG less often for medullary lesions 34.1% (95% CI: 25.9-42.3) than all other models 66.9% (95% CI: 62.2-71.6; p<0.001), despite this being a key site of NMOSD involvement.

**Figure 4.**
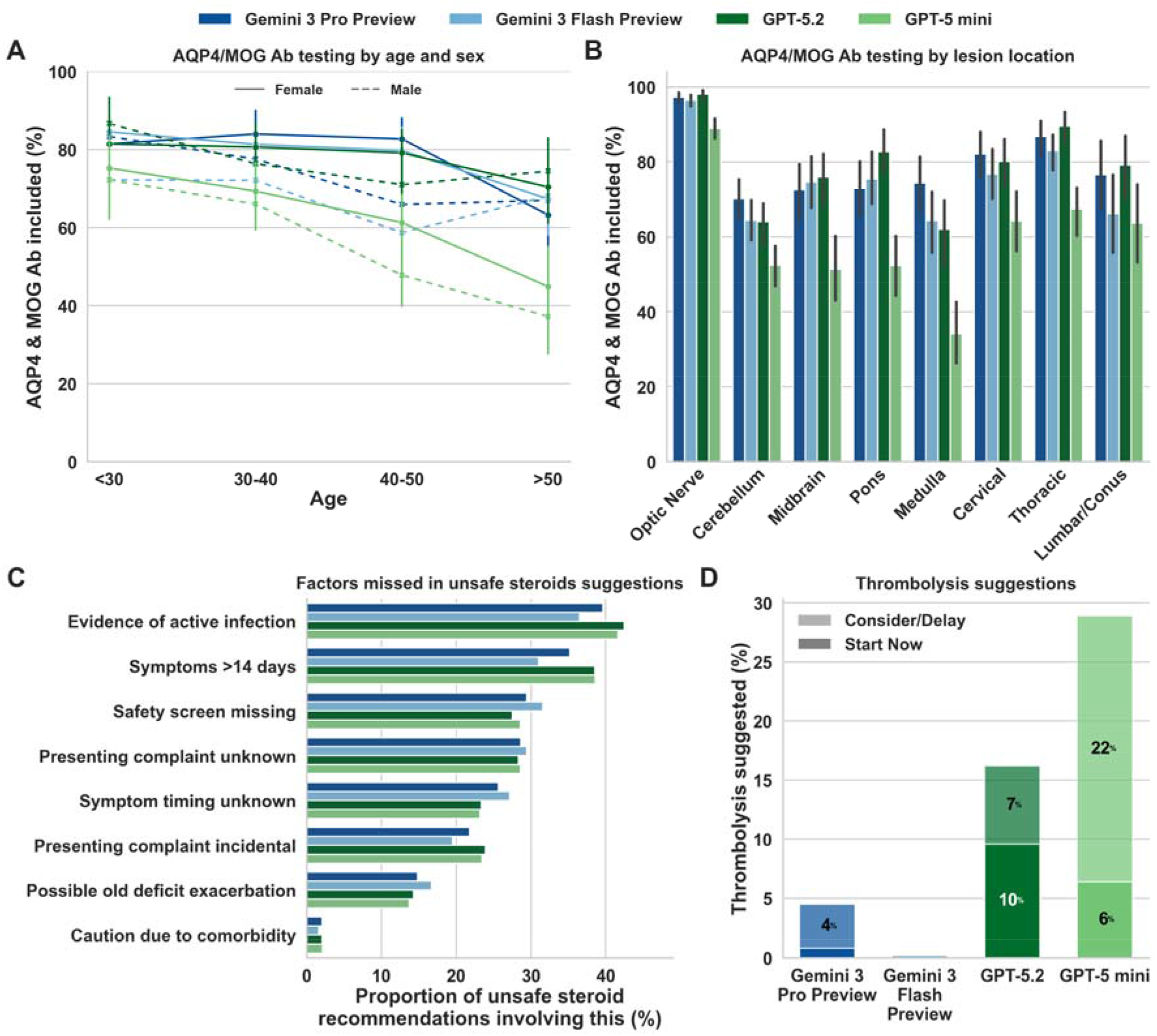
Impact of case-specific context upon LLM clinical decisions. **A)** Percentage of cases for which LLMs suggested testing AQP4 and MOG antibodies, categorised by patient age and sex. Model colours: Gemini 3 Pro Preview (dark blue), Gemini 3 Flash Preview (light blue), GPT-5.2 (dark green), and GPT-5 mini (light green). Solid lines represent female patients, and dashed lines represent male patients. **B)** Percentage of AQP4/MOG antibody testing suggestions across different lesion locations. **C)** Factors not acknowledged by LLMs in cases where they made potentially unsafe corticosteroid treatment recommendations. Each bar represents the percentage of unsafe decisions where a specific clinical safety concern, such as an active infection, or remote (>14 days since onset) symptom timing, was disregarded. **D)** Inappropriate thrombolysis recommendations. Stacked bars represent the frequency of recommendations for immediate (“Start Now”; solid/bottom) or delayed consideration (“Consider/Delay”; translucent/top) to initiate acute intravenous thrombolysis. Central values indicate the percentage of total cases for each recommendation type. Error bars in A/B represent standard error of the mean.

Analysis of management recommendations revealed consistent failures across all models. LLMs recommended high-dose steroids without acknowledging features that rendered them not indicated or contraindicated, such as explicit evidence of active infection, or when presenting complaints were incidental, confirmed as >14 days old, or lacked timing information (Figure 4C).

Strikingly, GPT-5.2 and GPT-5-mini recommended intravenous thrombolysis to “start now” in 9.6% (95% CI: 7.8-11.4) and 6.4% (4.9-7.9) of cases respectively, compared with <1% for both Gemini models (p<0.001) (Figure 4D).

Exploiting the advantages of our synthetic approach, we scaled the evaluation of GPT-5-mini to 10,000 cases. This allowed for a high-resolution analysis of rare edge-case behaviours such as the recommendation of intravenous thrombolysis for MS that would be statistically invisible in smaller, non-synthetic datasets (Figure 5A). The rate of immediate thrombolysis recommendations increased with patient age, from 3.9% (95% CI: 3.0-4.8) for patients <30 years to 6.8% (95% CI: 5.7-7.9) for >50 years (p=0.001, Cramer’s V=0.041).

**Figure 5.**
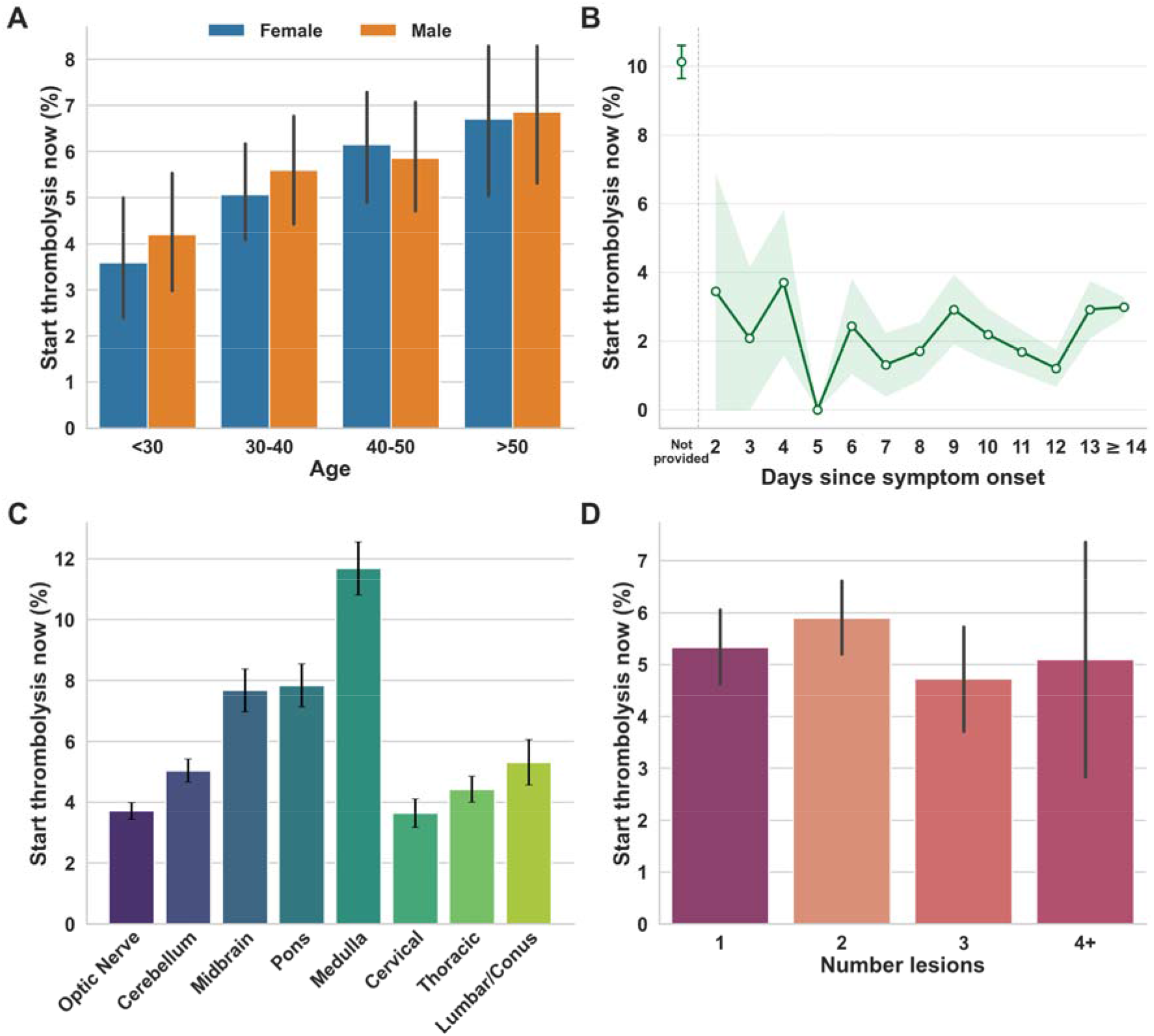
Detailed characterisation of inappropriate thrombolysis recommendations by GPT-5 mini (n=10,000). This analysis identified clinical factors associated with inappropriate “Start Now” thrombolysis decisions. **A)** Frequency of errors by patient demographics (blue = female; orange = male. **B)** Impact of whether presenting complaint timing was provided, and if so how many days since their onset. **C)** Error frequency by lesion location. **D)** Frequency of errors relative to the total number of ground-truth lesions. All error bars represent standard error of the mean (SEM).

Symptom timing influenced LLM recommendations more than whether presenting complaints were incidental or related to MS (Cramer’s V 0.162 vs 0.101 respectively). When symptom timing was not provided, thrombolysis recommendations reached 10.1% (95% CI: 9.2–11.1; Figure 5B). However, even with explicit description of an incidental presenting complaint starting >14 days ago, immediate thrombolysis was still recommended in 2.9% (95% CI: 1.6–4.2) of cases.

Medullary lesions were more frequently associated with immediate thrombolysis recommendations (11.7% [10.0–13.4] vs 4.9% [4.6–5.3] for all other regions; p<0.001; Figure 5C). Multifocal involvement across four or more disparate territories would be atypical for acute ischemia. However, total lesion burden had a negligible effect on thrombolysis recommendations (Cramer’s V=0.061; Figure 5D).

## DISCUSSION

At the surface level, modern frontier LLMs processed complex clinical information well, correctly including MS within differential diagnoses for most cases. However, our large-scale evaluation proves we cannot rely on assumptions of safety or broad, untargeted benchmarks to support AI use in healthcare^15–18^. Moving beyond aggregate benchmarks, our high-resolution approach reveals that superficial proficiency conceals unpredictable and catastrophic flaws in frontier AI clinical reasoning^9^. For instance, models routinely recommended immediate stroke thrombolysis for patients with MS and weeks-old, incidental complaints. We must therefore evaluate model behaviour across the full spectrum of diverse clinical presentations, a scale only possible with the automated, expert-validated approaches such as that introduced here.

This study was not designed to ‘rank’ models, nor do these LLMs make explicit clinical claims. However, our findings clearly indicate there is no universally ‘best’ LLM for clinical safety; performance was highly context-dependent^25^. High performance on superficial diagnostic tasks did not ensure safe decision-making, nor did a model’s ‘frontier’ status guarantee reliability. GPT-5-mini demonstrated safer steroid recommendations than the ostensibly ‘superior’ Gemini 3 Pro Preview, while Gemini models largely avoided the inappropriate thrombolysis suggested by GPT-5.2/mini. Conversely, some systemic deficits spanned all models, such as a universal inability to precisely localise spinal cord lesions. This pervasive imprecision persisted regardless of confounding factors such as the presence of multiple lesions causing additional long-tract signs, and perhaps reflects broader underrepresentation of granular spinal neuroanatomy in LLM training processes.

While obvious errors like inappropriate thrombolysis would likely be caught by human oversight, subtle blind-spots pose an arguably greater risk. One model’s systematic failure to test anti-AQP4/MOG for medullary lesions, a hallmark of NMOSDs^26^, illustrates this danger. Such specific omissions are invisible to standard benchmarks and easily missed by non-experts, which could lead to population-scale mismanagement. Massive clinical simulation is the only way to map the entire landscape of these edge cases and identify hidden deficiencies before real-world deployment.

In clinical practice, excellence also requires judiciousness, not a 100% action rate. For example, AQP4/MOG testing is only recommended when pre-test probability is high, to avoid false positives^27,28^. Human clinicians naturally vary in testing thresholds based upon individual preference, local protocols, or specific patient nuances. To integrate LLMs effectively we should be able to calibrate them to these nuances. Medical AI evaluation must move beyond simplistic ‘right or wrong’ grading to assess whether a model’s reasoning follows a logical clinical arc.

Current medical AI evaluation is hindered by slow manual review, shallow benchmarks, and the limitations of retrospective data (which lacks absolute ground truth and risks training-data contamination^22,23^). In contrast, our synthetic case generation executes in seconds. From 10,000 unique cases, we assessed 606,349 LLM clinical judgments. If a human expert took just 5 seconds per judgement, this scale would require 21 weeks of continuous expert labour; our automated system provided expert-level interrogation in hours. The only constraint to expanding across millions of scenarios was commercial application programming interface costs.

This work has limitations. First, synthetic cases, while diverse and realistic, may not perfectly replicate the full spectrum of real-world cases ^8,9^. Second, while we intentionally examined MS in depth, AI errors may differ across conditions. Our underlying system is expandable and capable of modelling most focal neurological and headache disorders^13^, but further expansion will be important. Third, we used multimodal inputs. However, LLM rationales accurately extracted image-embedded text (e.g. mentioning symptom durations) but failed during downstream synthesis (e.g. confabulating evidence). This indicates genuine deficits in higher-order reasoning, rather than image-parsing failures.

Our findings demonstrate the unprecedented insight afforded by automated, expert-level AI evaluation at scale. Traditional validation, relying on ‘typical’ cases and/or time-constrained manual human review, fails to capture the complex ‘edge cases’ where patient risk is greatest. We propose that independent, rigorous stress-testing across thousands of diverse scenarios become a prerequisite for deployment. Our methodology generates more unique clinical presentations than a clinician encounters in a lifetime^29^. Rather than just detecting failures, it provides the resolution to characterise the clinical drivers behind them and enable targeted guardrails before models expose patients to risk.

## Data Availability

The results, processed data and analysis code supporting the findings of this study will be made freely available upon publication at https://github.com/stepdaug. However, the raw case generation code and exact probability distributions used to define the simulated cohorts are not publicly available. These restrictions are in place to preserve the integrity of the simulation engine as a rigorous, future-proof evaluation tool for frontier medical AI. Public release of these datasets would risk contamination of future model training sets, thereby compromising the tools utility as a neutral benchmark.

## Data and code availability

The results, processed data and analysis code supporting the findings of this study will be made freely available upon publication at https://github.com/stepdaug. However, the raw case generation code and exact probability distributions used to define the simulated cohorts are not publicly available. These restrictions are in place to preserve the integrity of the simulation engine as a rigorous, future-proof evaluation tool for frontier medical AI. Public release of these datasets would risk contamination of future model training sets, thereby compromising the tool’s utility as a neutral benchmark.

## Acknowledgements

SDA is supported by a UK National Institute for Health and Care Research (NIHR) Clinical Lectureship and acknowledges infrastructure support for this research from the NIHR

Imperial Biomedical Research Centre (BRC). JV and MH received no specific funding for this work. GS, NIHR Advanced Fellow, NIHR302971 is funded by the NIHR for this research project. The views expressed in this publication are those of the author and not necessarily those of the NIHR, NHS or the UK Department of Health and Social Care. The authors also acknowledge MRC equipment support via the UK Dementia Research Institute and are grateful to Payam Barnaghi for assistance in accessing this.

## Author Contributions

SDA conception, design, case generation, data analysis, manuscript first draft and review. JV case validation and manuscript review. MH case validation and manuscript review. GS design and manuscript review.

