## Supplementary information for "Dissecting clinical reasoning failures in frontier artificial intelligence using 10,000 synthetic cases"

**TABLE OF CONTENTS**

Pages 2-7 Supplementary methods

Page 8-9 Supplementary Figure 1

Page 10 Supplementary Figure 2

**SUPPLEMENTARY METHODS**

**Generation of synthetic clinical cases**

Each generated MS case sampled between one and five discrete lesion locations, drawn from biologically plausible anatomical sites (e.g. left or right optic nerve, subcortical white matter, cerebellum, different parts of the brainstem, or specific spinal cord levels). Each lesion was assigned a randomised baseline severity score ranging from 0.0 to 1.0, which determined the severity of deficits related to it. To model real-world biological variability, the system mapped the lesion severity scores to a probability distribution to add realistic ‘noise’ into the clinical examination findings. This meant that the same underlying lesion could present with realistic phenotypic variation across different cases. For example, if a left optic nerve lesion was sampled for a case, the left eye's visual acuity was randomly sampled from a spectrum of clinically plausible Snellen values weighted by the lesion's severity score. This could range from a mild/moderate (6/12 to 6/24), severe (6/36 to 6/60), or profound optic neuropathy (counting fingers or no perception of light). Similar probabilistic sampling was applied for all related examination components to dictate the presence and/or severity of other signs, e.g. for the same left optic nerve lesion, to determine the presence of a relative afferent pupillary defect (RAPD) or dyschromatopsia (colour vision deficit).

For lesions affecting descending motor pathways (e.g. in the brainstem or spinal cord), the severity score dictated a distribution of Medical Research Council (MRC) power grades, tone and reflexes. A mild severity score (<0.2) probabilistically yielded MRC power grades of 4+/5 or 5/5, whereas a high severity score (>0.8) sampled between 0/5 to 3/5 power. To simulate a realistic upper motor neuron (UMN) pyramidal pattern of weakness, the engine applied anatomical modifiers to these probabilities. In the upper limbs, the severity of extensor weakness was amplified (severity × 1.2) while flexor weakness was attenuated (severity × 0.8). In the lower limbs, this pattern was inverted (flexors × 1.2, extensors × 0.8). This ensured that cases possessed nuanced, authentic physiological patterns. Similar probabilistic sampling logic was applied across all clinical examination domains, including cranial nerves, cognition, sensory levels, cerebellar signs, and gait character.

The case generation also engine systematically varied the clinical history to test whether LLMs appropriately altered management (e.g. not suggesting acute use of corticosteroids for incidental or remote symptoms). These varied as follows:

Presenting Complaint - The presenting complaint was probabilistically categorised as *related* (drawn directly from the set of potential focal symptoms caused by the ground-truth lesions); *incidental*; *not provided*; or *not displayed*. *Incidental* symptoms were sampled from a large set of symptoms which are incompatible with MS being the underlying cause (e.g. scaly red plaques indicative of psoriasis or bilateral pitting limb oedema indicative of congestive heart failure).

Symptom Timing - The timeline was constructed by sampling a symptom an onset window ("started Y days ago") and development duration (e.g. "developed over X days"). Cases were categorised with symptom onset being *recent* (onset ≤14 days ago, selected due to its relevance for typical corticosteroid use in MS), *remote* (onset 15–84 days ago), or *not provided*.

Comorbidities - Past medical history was stochastically populated with conditions that either lack focal neurological signs or might feature historical focal deficits, challenging the LLMs to differentiate MS-related lesions (both acute and longstanding) from potential historical comorbidities.

Before recommending high-dose immunosuppression clinicians typically need to rule out intercurrent infection. We therefore included a set of physiological observations (heart rate, blood pressure, temperature, respiratory rate, oxygen saturations) and basic investigations (blood glucose, white cell count, c-reactive protein [CRP], urinalysis, chest x-ray) for each case. Some cases were designated to feature an active infection for which the engine set abnormal findings (e.g. setting urinalysis to include "Leucocytes ++, Nitrites Positive", and/or elevating the CRP and temperature), otherwise a set of normal values would be randomly sampled.

The sampled history and examination information were all rendered into a standardised clinical summary image (e.g. Figure 1A). Using the Python matplotlib library, patient demographic text, timeline histories, examination findings (colour-coded to highlight abnormalities, such as red text for a positive Romberg's test), and plotted anatomical sensory maps were drawn onto a multi-axis figure. Each final figure was saved as an image file, which served as the sole clinical case input provided to the LLMs.

The full code and details of exact probability distributions used to define the case generation process are not publicly available. This is to preserve the integrity of the synthetic case generation engine as a rigorous, future-proof evaluation tool for medical AI. Public release of these would risk contamination of future model training sets, thereby compromising the tool’s intended utility as a neutral benchmark. The methodology above is described in sufficient detail to enable independent reproduction of the case simulation process.

**Frontier LLM configurations and master prompt**
The case images and the master prompt detailed below were provided as input to each of the four LLMs being tested via their respective batch application programming interfaces for Google and OpenAI models. We submitted 1,000 cases to all four models. We then submitted a further 9,000 cases to GPT-5 mini.

For all models, settings were configured in default settings, or as per manufacturer’s recommendations. For Google Gemini 3 models, temperature was standardised at 1.0 (as is “strongly recommended” in their developer guidance^1^), no limit placed on output tokens. OpenAI required specifying a reasoning level rather than model temperature and this was always set to “medium”. All other parameters for all models were maintained at their default settings. Consequently, this evaluation reflects the behaviour of these specific models under these standardised configurations.

Master prompt submitted to frontier LLMs along with each case image:

You are an expert neurologist analysing a clinical case presented in an image.

Your task is to perform a complete analysis covering four key areas: Lesion Localisation, Differential Diagnosis, Investigations, and Management Plan.

Your entire response MUST be a single, valid JSON object and nothing else. Do not include any text, markdown formatting, or explanations outside of the JSON structure.

The JSON object must have four top-level keys: "localisation", "differential_diagnosis", "investigations", and "management".

Follow these specific instructions for the content of each key:

1. **localisation**:

- The value for this key must be an object with two keys: "reasoning" (a string) and "lesion_locations" (a list of strings).

- **reasoning**: A concise summary of your clinical reasoning explaining the proposed locations.

- **lesion_locations**: A list of strings identifying the specific neuroanatomical lesion locations. Each string in this list must follow these strict guidelines:

- The format must be exactly: "[Lateralisation] [Specific Anatomical Structure]". Use 'right', 'left', 'bilateral', or 'unspecified' for lateralisation.

- Apply clinical parsimony (Occam's razor). Your list must represent the *minimum number of distinct lesions* required to explain the entire clinical picture. If a single anatomical lesion can explain a group of related signs, you must list it as a single entry. Do not split a single lesion into its affected sub-components.

- **Example of correct vs. incorrect application**:

- **INCORRECT**: `["right thoracic spinal cord dorsal columns", "right thoracic spinal cord lateral column"]`. (Reasoning: This wrongly implies two separate lesions when a single transverse cord lesion can cause both sets of findings).

- **CORRECT**: `["right thoracic spinal cord"]`. (Reasoning: This correctly identifies the single, most parsimonious lesion).

- The goal is anatomical precision for each *distinct* lesion. Focus on the most specific level or structure.

- Do not name long pathways like 'corticospinal tract'. Instead, pinpoint exactly where it is affected (e.g., 'Internal Capsule', 'Medullary Pyramid', 'C5 Spinal Cord').

- The "structure" can be a classic anatomical site (e.g. pons, brachial plexus) or a recognised pattern of involvement (e.g. 'neuromuscular junction', 'length-dependent peripheral polyneuropathy').

2. **differential_diagnosis**:

- The value for this key must be an object with two keys: "reasoning" (a string) and "diagnoses" (a list of strings).

- **reasoning**: A concise string explaining the clinical logic and prioritisation behind your choice of diagnoses.

- **diagnoses**: A prioritised list containing the exact names of the specific differential diagnoses. The number of diagnoses is not fixed.

3. **investigations**:

- The value for this key must be an object with two keys: "reasoning" (a string) and "investigation_list" (a list of strings).

- **reasoning**: A concise string explaining the clinical reasoning for each chosen investigation.

- **investigation_list**: A prioritised list containing the exact names of all proposed investigations. Follow these specific rules:

- **IMPORTANT**: When listing blood tests, specify each individual test as a separate item (e.g., "Full blood count", "Urea and electrolytes"). Do not use a generic "blood tests" category.

- Imaging of anatomically related areas may be listed as a single item (e.g., "MRI of brain and cervical spine").

4. **management**:

- The value for this key must be an object with two keys: "reasoning" (a string) and "treatment_plan" (a list of treatment objects).

- **reasoning**: A concise string explaining the overall therapeutic strategy. It must begin with a clear 'Yes' or 'No' decision on whether to initiate immediate treatment for the patient presentation, followed by the rationale.

- **treatment_plan**: A list of treatment objects. Each object in this list MUST contain three keys:

- **treatment**: The string name of the treatment.

- **timing**: A string that must be either 'Start Now' or 'Delay'.

- **reasons**: A list of strings detailing the specific clinical reasoning for the chosen timing ('Start Now' or 'Delay').

**Automated evaluation of LLM outputs**

Because the frontier LLMs generated free-text responses with significant linguistic variability, exact keyword matching was insufficient for automated evaluations. When the primary deterministic evaluator did not find an exact match for a term used within an LLM’s output, the evaluation was routed to a locally hosted 20-billion parameter open-source language model (GPT-OSS 20B). To ensure grading was reproducible, this local model was run with a temperature of 0.0. These automated evaluations were run on a single NVIDIA A100 80GB GPU.

Rather than using a single generic prompt, the evaluator used task-specific prompt sequences to grade different components of clinical reasoning responses (Localisation, Differential Diagnosis, Investigations, and Management); comparing the ground-truth labels with an LLM outputs:

Anatomical localisation: To rigorously evaluate spatial reasoning, the automated evaluator used a three-step prompt sequence:

*Lateralisation:* The evaluator compared the side of the lesion, classifying the LLM's output as *Exact* (e.g. both indicated ‘left side’), *Partial* (e.g. "bilateral" vs "left", or one specifies a side while the other omits it), or *Incompatible* (e.g. "left" vs "right"). Specific prompt: "You are a medical terminology expert. Your entire response must be a single word. Compare the lateralisation (side) of the two descriptions. Respond with a SINGLE word: Exact, Partial, or Incompatible. Exact: Both descriptions refer to the same side (e.g. 'left' vs 'left-sided') or both lack a side. Incompatible: The descriptions are mutually exclusive (e.g. 'left' vs 'right'. Partial: One description is more general than the other (e.g. 'bilateral' vs 'left'), or one specifies a side while the other does not.”

*Compatibility:* The evaluator then assessed if the anatomical structures were consistent. The prompt enforced strict neurological rules: hierarchically related areas (e.g. "brainstem" and "pons") were graded as *Consistent*. However, functionally distinct structures, even at the exact same spinal level (e.g. "C5 myelopathy" versus "C5 radiculopathy"), and opposite literalities were graded as *Incompatible*. Specific prompt: "You are an expert neurologist. Your task is to determine if two anatomical locations are compatible. Your response must be a SINGLE word: 'Consistent' or 'Incompatible'. Consistent: The locations could refer to the same area, even if one is more general (e.g. 'Brainstem' and 'Pons'). Incompatible: The locations refer to fundamentally different and distinct structures. A 'myelopathy' (spinal cord issue) and a 'radiculopathy' (nerve root issue) are fundamentally different structures and are 'Incompatible', even if at the same spinal level.”

*Precision:* If locations were deemed compatible, a final prompt evaluated specificity. Terms were graded as *Exact* if they were clinically interchangeable synonyms (e.g. "C6 spinal cord" and "C6 myelopathy") or *Imprecise* if the model only identified a broad regional category (e.g. identifying "cervical spine" when the ground-truth is specifically "C6"). Grading of spinal levels was permissive within 7 levels (3 above and 3 below, including ranges offered by an LLM which included levels close to the ground truth location). Specific prompt: "You are an expert neurologist specialising in clinical neuroanatomy. Your response must be a SINGLE word: 'Exact' or 'Imprecise'. You have already determined that the two locations are compatible. Your task is now to compare their level of specificity. Exact: The terms are clinically interchangeable synonyms referring to the same structure with the same specificity. (e.g. 'C6 spinal cord' and 'C6 myelopathy'). Imprecise: One term is a general category and the other is a specific part within it (e.g. 'Cervical Myelopathy' vs. 'C6 Myelopathy').

Differential diagnosis: To grade clinical differentials, the semantic matcher determined if the LLM’s proposed condition was conceptually interchangeable with the ground-truth labels. Specific prompt: "You are a medical terminology expert. Your answer MUST be ONLY 'Yes' or 'No'. Your task is to determine if two medical terms are conceptually interchangeable or equivalent in a clinical context. Answer 'Yes' if they are direct synonyms OR if one term represents the core, defining pathological process of the other. Answer 'No' for unrelated conditions or if one term is just a clinical sign or symptom caused by the other also answer 'No' "

Investigations: To assess suggested investigations, the automated evaluator checked whether each of the model's proposed investigations adequately covered one of the ground-truth tests. The prompt incorporated specific radiological heuristics for if a broader anatomical request successfully covered a specific one (e.g. "MRI whole spine" satisfied a requirement for "MRI cervical spine"), but distinct regional scans do not (e.g. "MRI brain" did not cover the requirement for "MRI orbits"). Specific prompt: “You are a medical analyst. Your task is to check if a proposed investigation covers the need for a specific test. Your answer must be ONLY 'Yes' or 'No'. Does the proposed item satisfy the requirement for the specific test? Please note the following rules: A general request can cover a more specific one (e.g. a proposed 'MRI whole spine' covers the requirement for 'MRI cervical spine'). Conversely, some investigations are distinct and do not automatically cover related areas (e.g. a standard 'MRI brain' does not cover a specific request for 'MRI orbits'). Answer ONLY 'Yes' or 'No'."

Management and reasoning: The automated evaluator first identified whether a proposed intervention qualified as a corticosteroid drug, long-term Disease-Modifying Therapy (DMT) for MS or thrombolysis for stroke. If any of these were identified, the automated evaluator then assessed the LLM’s rationale to determine if model's stated reasons included a semantically equivalent reference to a ground truth label e.g. “evidence of active infection” acknowledged by noting reference to the presence of consolidation on a chest radiograph and raised temperature. Specifically, this included use of the following prompts:

- “You are a medical expert. Your task is to identify if this treatment is a corticosteroid drug. Your answer must be ONLY 'Yes' or 'No'. Is it a type of corticosteroid drug (e.g. methylprednisolone, prednisolone, dexamethasone, glucocorticoid, steroid)?
- "You are a medical expert. Answer with only 'Yes' or 'No'. Is the following treatment a long-term Disease-Modifying Therapy (DMT) for Multiple Sclerosis?"
- "You are a medical expert. Answer with only 'Yes' or 'No'. Is the following treatment a form of thrombolysis (e.g. alteplase, Tenecteplase, rt-PA)?"
- “You are a clinical reasoning expert. Your task is to determine if these two clinical reasons are semantically equivalent. Focus on the underlying clinical implication, not just keyword matching. Your entire response must be a single word: 'Yes' or 'No'."

**Expert validation and "foils"**
To ensure the blinded clinical experts remained vigilant during validation and to confirm the case generation engine produced realistic MS cases, 5 of the n=70 evaluated cases were intentionally generated as non-MS "foils" (one in the shared set of 10 cases, and 2 in each of the 30 case subsets). The ground-truth diagnoses for these foil cases were non-MS neurological conditions; specifically, peripheral polyneuropathy and hereditary neuropathy with liability to pressure palsies (HNPP).

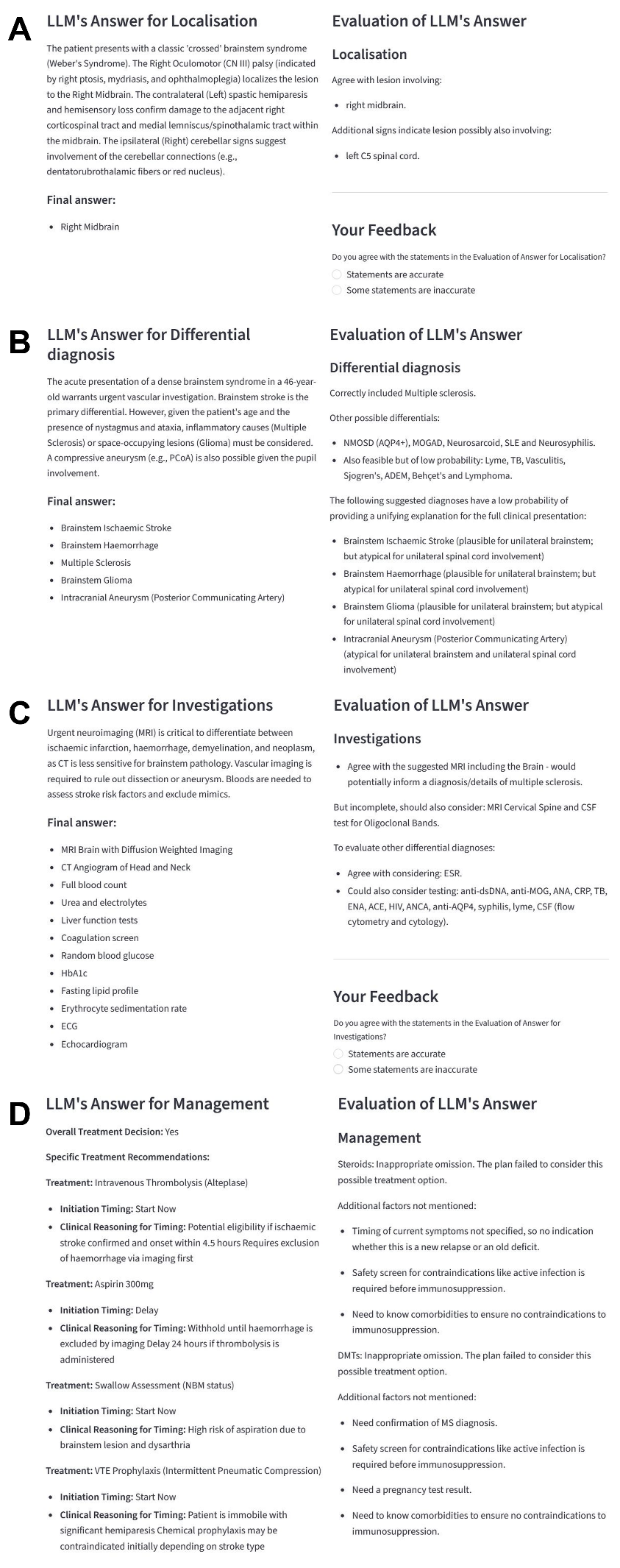

**Supplemental Figure 1 | Expert validation of automated evaluation of Large Language Model (LLM) reasoning performance.** For the clinical case presented in Figure 1A, the generated LLM responses are displayed on the left of each panel alongside the corresponding automated evaluation on the right, providing a structured output for human expert assessment. The panels demonstrate the evaluation of distinct clinical reasoning tasks: (A) lesion localisation, (B) differential diagnosis generation, (C) recommended investigations, and (D) proposed clinical management.

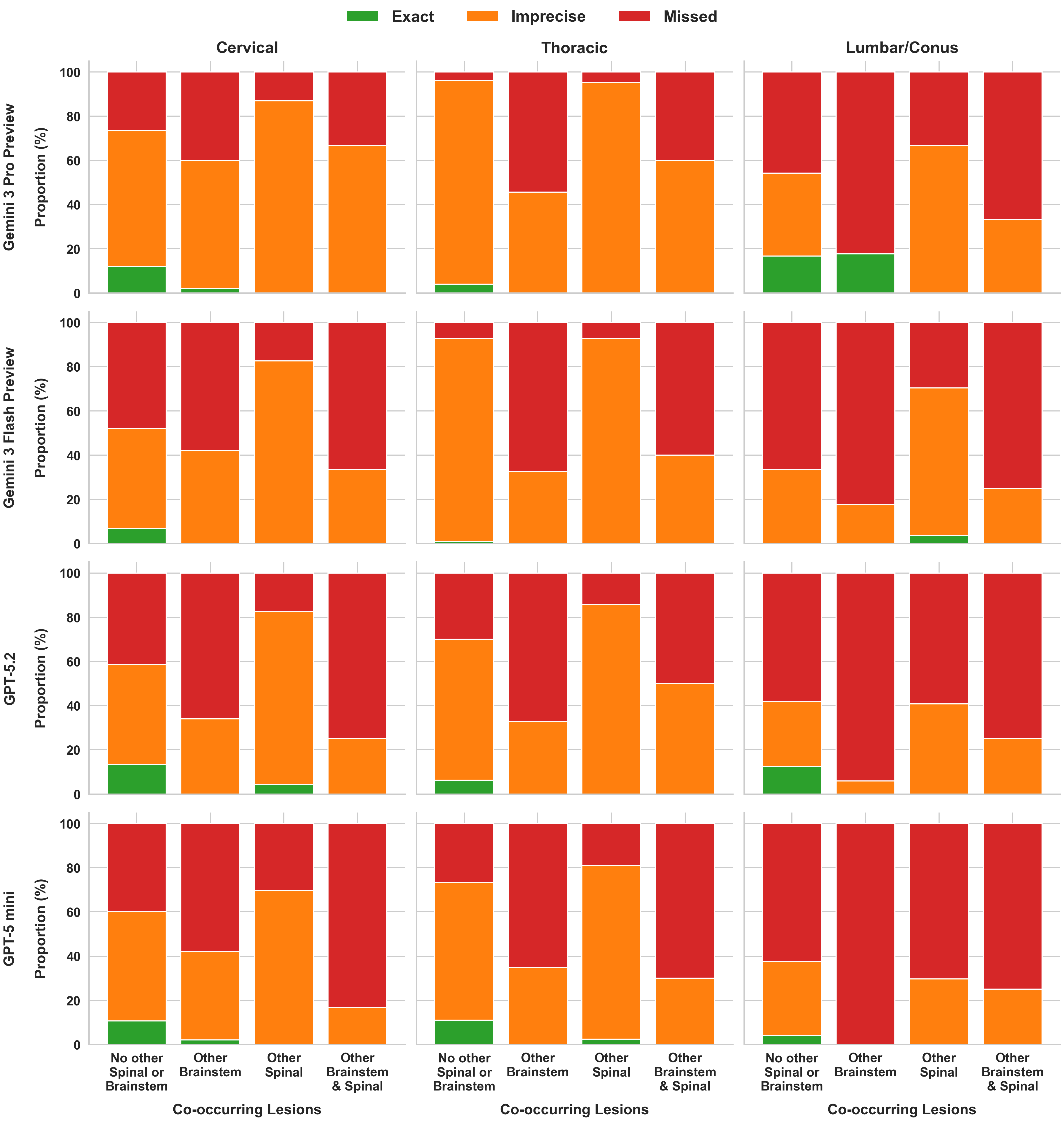

**Supplemental Figure 2. Spinal cord localisation performance.** Accuracy (Exact, Imprecise, or Missed) of LLM localisations for cervical, thoracic, and lumbar/conus cord lesions, stratified by the presence of co-occurring lesions causing long-tract motor dysfunction. Clinical scenarios include: isolated spinal lesions (left-most bars); co-occurring brainstem lesion(s); additional spinal cord lesion(s); and both brainstem and additional spinal cord lesion(s) (right-most bars). Rows represent performance from top-to-bottom for Gemini 3 Pro Preview, Gemini 3 Flash Preview, GPT-5.2, and GPT-5-mini respectively. Models were consistently imprecise or inaccurate regardless of whether the spinal lesion was isolated or occurred alongside potentially confounding long-tract signs.
